# “*Creating a culturally competent pharmacy profession*”: a qualitative exploration of pharmacy staff perspectives of cultural competence and its training in community pharmacy settings

**DOI:** 10.1101/2023.03.08.23286977

**Authors:** Jessica McCann, Wing Man Lau, Andy Husband, Adam Todd, Laura Sile, Amandeep Kaur Doll, Sneha Varia, Anna Robinson-Barella

## Abstract

**Introduction:** Cultural competence is an important attribute underpinning interactions between healthcare professionals, such as pharmacists, and patients from ethnic minority communities. Health- and medicines-related inequalities affecting people from underrepresented ethnic groups, such as poorer access to healthcare services and poorer overall treatment outcomes in comparison to their White counterparts, have been widely discussed in existing literature. Community pharmacies are a first port of call healthcare service accessed by diverse patient populations; yet, limited research exists which explores the perceptions of culturally competent care within the profession, or the delivery of cultural competence training to community pharmacy staff. This research seeks to gather perspectives of community pharmacy teams relating to cultural competence and identify possible approaches for the adoption of cultural competence training.

**Methods:** Semi-structured interviews were conducted in-person, over the telephone or *via* video call, between October-December 2022. Perspectives on cultural competence and training were discussed. Interviews were audio-recorded and transcribed verbatim. Reflexive thematic analysis enabled the development of themes. QSR NVivo (Version 12) facilitated data management. Ethical approval was obtained from the [anonymised] University Ethics Committee (ref: 25680/2022).

**Results:** Fourteen participants working in community pharmacies were interviewed, including: 8 qualified pharmacists, 1 foundation trainee pharmacist, 3 pharmacy technicians/dispensers and 2 counter assistants. Three themes were developed from the data which centred on: (1) defining and appreciating cultural competency within pharmacy services; (2) identifying pharmacies as “cultural hubs” for members of the diverse, local community; and (3) delivering cultural competence training for the pharmacy profession.

**Conclusion:** The results of this study offer new insights and suggestions on the delivery of cultural competence training to community pharmacy staff, students and trainees entering the profession. Collaborative co-design approaches between patients and pharmacy staff could enable improved design, implementation and delivery of culturally competent pharmacy services.

**Patient or public contribution:** The Patient and Public Involvement and Engagement group at [anonymised] University had input in the study design and conceptualisation. Two patient champions inputted to ensure that the study was conducted, and the findings were reported, with cultural sensitivity.

**Trial registration:** Not applicable. Ethical approval was obtained from the [anonymised] University Research and Ethics Committee (reference: 25680/2022).

## Introduction

Cultural competence can be defined as an individual’s ability to possess the skills and knowledge to effectively interact with people from different cultural backgrounds.^1^ It involves the acknowledgement, understanding and appreciation of an individual’s cultural identity such as their religion, ethnicity, nationality, gender and sexual orientation.^2,3^ Evidence has demonstrated that being culturally competent promotes communication between individuals,^4^ respect for other cultures, individual self-awareness^1^ and support shared decision-making between individuals.^5^ Therefore, it can be considered a key attribute to those working within the healthcare system.^6^

The General Pharmaceutical Council Standards for Pharmacy Professionals report demonstrates the responsibility of pharmacy professionals to ‘*treat people as equals, with dignity and respect, and meet their own legal responsibilities under equality and human rights legislation, while respecting diversity and cultural differences*… *(and) assess and respond to the person’s particular health risks, taking account of individuals’ protected characteristics and background”*.^*7*^ Similar statements are acknowledged in the General Medical Council (GMC) Equality, Diversity and Inclusion Policy, which ensures that the organisation and all medical professionals *“treat anyone who [we] interact with fairly, without bias or discrimination*”,^8^ and in the principles of good practice for community pharmacy teams, to address health inequalities (point 1.2.6) in the National Institute of Health and Care Excellence Guidance.^9^ Despite referring to cultural competence within the guidance from professional bodies, there remains evidence of healthcare and medicines inequalities affecting patients belonging to ethnic minority communities.^5,10,11^

Recent studies have identified several factors which may contribute to these health inequalities, including poorer health outcomes, lower reported health literacy levels, lower socio-economic status, and feelings of disempowerment and distrust within the healthcare system for those from underrepresented ethnic communities, compared to their White counterparts.^12,13^ Evidence suggests that one approach of tackling the aforementioned health inequalities could relate to the education and training of healthcare professionals, particularly developing skills to become culturally competent.^11,14-16^ Govere *et al*. demonstrated that cultural competence training had a positive impact on cultural awareness and overall competence of healthcare professional consultations, hence improving rates of patient satisfaction.^14^

Most knowledge around training of cultural competency within healthcare settings currently focuses on the fields of nursing, medicine and dentistry. There have been variations in proposed strategies and frameworks for teaching cultural competence to these healthcare professionals;^17^ for example, including the provision of online, self-directed learning sessions for trainees,^18^ as well as face-to-face workshops and seminars, delivered by trainers.^19-22^ One setting within healthcare that encounters a wide range of culturally diverse patients is community pharmacy. Community pharmacy is regarded as a vital and easily accessible healthcare setting to any patient who requires health advice and treatment.^23^ A recent study proposed that cultural competence training should be implemented into the training curriculum of all staff working within community pharmacies;^10^ however, limited research exists on the optimal delivery methods of cultural competence training to meet this need.^24,25^

By exploring the perspectives of community pharmacy staff members, this qualitative study aims to: (i) provide new insights that showcase beliefs and attitudes towards cultural competence within the pharmacy profession and (ii) identify strategies to implement and deliver training for community pharmacy staff to use within their culturally diverse places of work.

## Method

### Recruitment and sampling

The consolidated criteria for reporting qualitative research (COREQ) checklist was followed for this work (see Multimedia Appendix).^26^ Given the capabilities of digital strategies to support qualitative research, a blended-strategy was applied to perform participant recruitment and data collection with pragmatism. Recruitment was facilitated by community pharmacies, community charities and professional networks based in the North East of England, as well as on the social media profiles of two members of the research team (JMcC and AR-B). All interested participants who contacted the research team were emailed an information sheet and consent form detailing the purpose and aim of the research. Those who expressed an interest and gave their written consent, were enrolled to the study. There was no relationship established between the researcher and participants prior to study commencement or recruitment. Inclusion criteria comprised: participants over 18-years of age who held a role within a community pharmacy team working in the UK (including, but not limited, to: pharmacists, foundation trainee pharmacists, pharmacy technicians, dispensers/dispensing staff and counter assistants). Purposive sampling was used to recruit participants and ensure representation from a variation of typical job roles within community pharmacy teams; participants were also of mixed age ranges, had been qualified in their job role for varying lengths of time and were from varying ethnic backgrounds.

### Semi-structured interviews

In-depth, semi-structured interviews were conducted by one researcher (JMcC, a female pharmacy student researcher with experience of qualitative research) between October and December 2022. Interviews were conducted either remotely, *via* Zoom® or telephone calls, or in-person (face-to-face); all participants were offered the choice of which format they would prefer. The semi-structured interview topic guide (see Supplementary File) was developed based on three pilot interviews and covered key issues identified in the existing literature,^5,10,11^ including participants’ knowledge of cultural competence within pharmacy and wider healthcare settings; participants’ perspectives and experiences of interacting with patients from ethnic minority communities; and their views and suggestions on cultural competence training. In addition, the topic guide was informed by findings from a previous qualitative study conducted by the research team^5^ and the lived experiences of patient champions involved as co-authors in this study (LS and AKD).

### Data analysis

All semi-structured interviews were audio-recorded to enable data analysis. The audio files were encrypted and transcribed verbatim by JMcC; immediately following transcription, the audio files were deleted. All interview data were anonymised at the point of transcription and all transcripts were checked for accuracy and correctness by AR-B. Participants did not provide comment on the transcripts nor feedback on results.

Following reflexive thematic analysis processes, as defined by Braun and Clarke,^27,28^ the principle of constant comparison guided an iterative process of data collection and analysis. Reflexive thematic analysis was performed by two researchers (JMcC and AR-B) to analyse the interview data. Close and detailed reading of the transcripts allowed the two researchers to familiarise themselves with the data. Initial descriptive codes were identified in a systematic manner across the data sets; these were then sorted into common coding patterns, which enabled the development of analytic themes from the data. The themes were reviewed, refined and named once coherent and distinctive. Two authors (JMcC and AR-B) performed the data analysis through discussion and, if agreement was not reached, by consensus with the wider research team (LS, AKD, WML, AKH and AT). Post-interview field notes enhanced this reflective process. NVivo (version 12) software was used to facilitate data management. The research team were in agreement that data saturation (or data sufficiency and information power) occurred after conducting 12 semi-structured interviews and thus, study recruitment stopped following interview number 14.^29^ To ensure confidentiality when using direct patient quotes within this research, non-identifiable pseudonyms are used throughout, *e*.*g*., Participant 1 and Participant 2 *etc*.

### Considerations when reporting participant demographics

The researchers wished to consider whether there were any connections or associations between perspectives shared by participants, and their ethnicity. Collecting data on a person’s ethnic group is complex, since ethnicity in itself is a multi-faceted and changing phenomenon.^30^ Various ways of measuring ethnicity exist and could include a person’s country of birth, nationality, religion, culture, language, physical appearance or a combination of all of these aspects.^3,31^ Efforts have been taken in this study to report a multitude of these aspects, to demonstrate the multi-faceted layers that accompany discussions about ethnicity. The UK Office of National Statistics ‘Ethnic group, national identity and religion’,^3^ the UK Census Reporting Classification,^32^ and the National Institutes of Health (NIH) ‘Racial and Ethnic Categories and Definitions for NIH Diversity Programs and Other Reporting Purposes’^31^ guides were used to inform the reporting of participant ethnicity for this study (as demonstrated in Table 1). Table 1 also includes a column for self-identified ethnicity and is reported verbatim from each participant’s interview.

**Table 1:** Participant demographics

| Participant number | Reported sex | Age range (years) | Staff role within Community Pharmacy | Self-reported ethnicity | Interview format | Interview duration (minutes, seconds) | Region of practice within UK |
| --- | --- | --- | --- | --- | --- | --- | --- |
| 1 | Female | 20-29 | Pharmacist | White, British | Video-call (Zoom) | 42m 38s | Northumberland / Tyne & Wear |
| 2 | Female | 20-29 | Dispenser | White, British | In-person | 20m 19s | York |
| 3 | Female | 40-49 | Counter Assistant | Mixed-race, British | Telephone | 22m 07s | Northumberland |
| 4 | Female | 30-39 | Pharmacist | White, Scottish | Telephone | 39m 52s | Glasgow |
| 5 | Female | 40-49 | Pharmacist | White, British | Video-call (Zoom) | 52m 22s | Northumberland / Tyne & Wear |
| 6 | Male | 20-29 | Pharmacist | White, British | Video-call (Zoom) | 55m 19s | Northumberland / Tyne & Wear |
| 7 | Female | 30-39 | Pharmacist | White, British | Video-call (Zoom) | 41m 39s | Northumberland |
| 8 | Female | 40-49 | Pharmacy Technician | White, British | Video-call (Teams) | 26m 46s | London |
| 9 | Female | 30-39 | Pharmacist | Chinese | Video-call (Teams) | 32m 22s | Northumberland / Tyne & Wear |
| 10 | Male | 40-49 | Pharmacist | Pakistani | Video-call (Zoom) | 23m 21s | South Yorkshire |
| 11 | Male | 30-39 | Pharmacist | Chinese | Video-call (Zoom) | 39m 52s | York and London |
| 12 | Female | 20-29 | Counter Assistant | White, British | Telephone | 22m 41s | Northumberland |
| 13 | Male | 20-29 | Dispenser | White, British | Telephone | 25m 21s | Tyne & Wear |
| 14 | Female | 20-29 | Foundation Trainee Pharmacist | White, British | Video-call (Zoom) | 30m 04s | Tyne & Wear |

### Ethical approval

Ethical approval was obtained from the [anonymised] University Ethics Committee (reference: 25680/2022) and research governance was followed in accordance to [anonymised] University research policies.

## Results

### Participant demographics

Fourteen participants in total were recruited and interviewed for this study (participant characteristics are described in Table 1). Of the fourteen participants interviewed; eight described their job role within community pharmacy as pharmacists (57%), one interviewee was a foundation trainee pharmacist (8%), three were dispensers/pharmacy technicians (21%) and two were counter assistants (14%). Ten participants self-reported their ethnicity to be White, with nine stating they were British and one stating they were Scottish; one participant identified as mixed-race British; two participants identified as Chinese, and one identified as Pakistani. The average age of the participants was 30 years (SD ±8.06). Participants worked within community pharmacy settings across the UK; specifically, nine participants worked across the regions of Northumberland, Tyne and Wear (England), three participants worked across Yorkshire (England), two participants stated they worked in London (England), and one participant worked in Glasgow (Scotland). Nine interviews were conducted over video call-based software (64%), four interviews were conducted over the telephone (28%) and one interview was carried out in-person (8%). There were no refusals to partake, participant drop outs or repeat interviews.

Three overarching themes were developed to reflect the perceptions of community pharmacy staff on cultural competence and the delivery of cultural competence training within community pharmacy. These focused on (1) defining and appreciating cultural competency within pharmacy services; (2) identifying pharmacies as “cultural hubs” for members of the diverse, local community; and (3) delivering cultural competence training for the pharmacy profession (Figure 1). The three themes, and their sub-themes, are discussed in turn.

**Figure 1.**
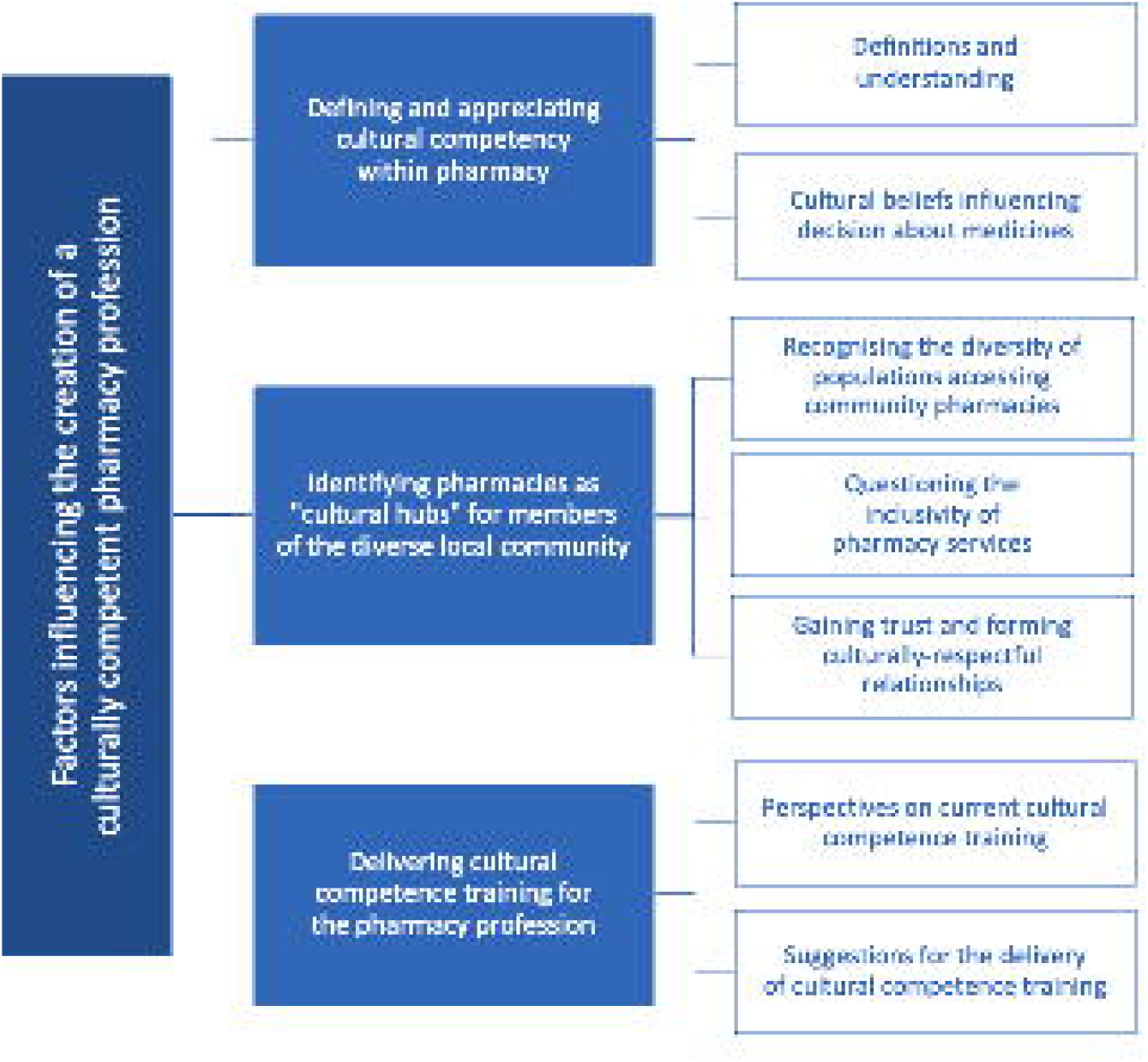
Factors influencing the creation of a culturally competent pharmacy profession: developed from the perspectives of pharmacy professionals

### Theme 1: Defining and appreciating cultural competency within pharmacy

#### Definitions and understanding

Cultural competence was believed by most participants to relate to the awareness and appreciation of another person’s culture, with one explaining that being culturally competent is *“about understanding and appreciating people from a lot of diverse backgrounds – that are different to yours. So that’s whatever ethnicity you are, it’s understanding about cultures and upbringing and all the things that make you, you”* (Participant 8). In this sense, cultural competence was interpreted, in a wider social context, as a skill that an individual can develop through interactions with people from different cultures. Other participants described cultural competence as a concept relative to health, and drew on their medicines expertise to address this; they described cultural competence as being *“inclusive in your treatment of those patients and being able to build that into the way we approach patient care and healthcare decisions”* (Participant 6).

> *“You’ve got to be aware of issues such as religion, race or cultural differences that may require a different approach to treating that patient*… *you know what they can and can’t eat for example, you can offer them alternative medication with products that wouldn’t be offensive or unacceptable to them” (Participant 5)*.

#### Cultural beliefs influencing decisions about medicines

Participants provided examples where it was important to be aware of formulation considerations and excipients within medications. Specifically, the suitability of capsules containing pork products for someone practicing religions including Islam or Judaism was discussed, with one participant stating *“somebody who’s Muslim wouldn’t be happy with pork products, like gelatine, in their capsules”* (Participant 4). Others also recalled examples of investigating alternative medications to suit a patient’s cultural needs. A pharmacist explained, *“it took me probably a couple of hours to source the medication in liquid form… I had to ring manufacturers and contact lots of different companies to try and find an alternative and (pause) it took a lot of time”* but they *“knew it was super important to them [the patient] and if was me in their shoes, I wouldn’t have taken that medication because it’s not acceptable”* (Participant 5). It was acknowledged by many participants that although it may take additional time to source suitable medications for patients, it was important for delivering high quality care, and there was an underpinning belief that these additional considerations would lead to a more individualised approach to patient care.

Other participants reflected that the beliefs of certain cultures may impact on an individual’s ability to engage with healthcare access or medicines-taking. It was suggested that certain cultural beliefs could pose as barriers to individuals accessing the care that they need, as there may be cultural stigma surrounding certain health conditions. A pharmacist recalled an example where a Muslim patient, who was receiving methadone treatment, said he would be *“mortified if a person from their community had seen them”* (Participant 4). The pharmacist contemplated that *“some people perceive that Muslims don’t drink and therefore it’s unusual for a Muslim to end up on methadone or a dependent substance”* (Participant 4). They hinted that patients should not be labelled under common stereotypes and that health professionals should know to provide equal treatment to patients, regardless of their cultural background.

> *“I think it’s about being concerned that they [patients] may be stigmatised within their community… or if it’s perhaps not something that’s seen as appropriate in their religion. It’s a little bit about shame, and personal shame and er, feeling like they’re going against the cultural norm for them. I think that stigma could potentially prevent them seeking help for their conditions” (Participant 4)*.

Two participants reflected on experiences of barriers that prevented a person seeking healthcare or medication advice, because they were *“a female (pharmacy professional) and their (the patient) religion means they can’t, or shouldn’t, interact with females”* (Participant 5). Suggestions were given around possible ways to overcome such barriers, even *“how pharmacy regulators could engage with key members of those communities to try and explain how we can work together to overcome that”* (Participant 5).

> *“I think that one of the hardest challenges is where maybe the gender of the person who delivering the care, is not seen as being appropriate to speak to a male member of the public and say, ‘It’s ok to engage with a female member of staff at the pharmacy. You’re not going to get in trouble if you do that’” (Participant 5)*.

### Theme 2: Identifying pharmacies as “cultural hubs” for members of the diverse local community

#### Recognising the diversity of populations accessing community pharmacies

Participants discussed that community pharmacies are unique healthcare settings; they are an accessible *“source of health advice and health care, where you can speak to a qualified professional, all without requiring an appointment”* (Participant 3). One participant reflected that being a community pharmacist meant *“you are often the first point of contact like in the community for any patient*” (Participant 1) and one counter assistant recognised the diversity of patients that attend community pharmacies, stating *“it’s not always the same ethnicity (within the patients we are treating) and you’re rarely going to get a pharmacy that only has one ethnicity (in its population)”* (Participant 12). Participant views across all job roles within community pharmacy described the potential for interactions with members of a diverse population accessing the pharmacy. One participant viewed the potential for community pharmacies to be regarded as *“cultural hubs”* that local communities can access, knowing they will *“be recognised and spoken to in way that accounts for them and their culture”* (Participant 11).

Most of the participants had experience working within various pharmacies across the UK and were able to recall differences in the level of diversity between the pharmacies on a national-level, as well as on a local-level. One participant discussed their experience of working as a locum pharmacist across a number of cities across the North East of England, where diversity differences were even noticed *“only 2-miles down the road you’ve got an entirely different community living there and they’ll most likely need different approaches to people living 2-miles the other direction”* (Participant 6). Another pharmacist acknowledged diversity differences comparing inner-city pharmacies; they described *“particularly in the North East, like in [name of major city], you can cross the bridge and find communities that are highly populated with Jewish people, or you can cross another bridge and find areas even more diverse again, like housing for refugees or asylum seekers from War torn countries”* (Participant 6). There was also suggestion that pharmacies located in areas of higher deprivation contained a greater population of ethnic minority patients. One pharmacist discussed noticing diversity variations between two pharmacies they worked at with *“different sort of socio-economic areas between [names of two villages]”* (Participant 7). As well, a participant who worked in Scotland suggested *“where I work, [name of city], it’s like an affluent rural area… so I would say predominantly the community is Caucasian, middle class people”* (Participant 4).

#### Questioning the inclusivity of pharmacy services

There was variation between participant perspectives on whether their pharmacy services demonstrated inclusivity towards patients from ethnic minority groups. Most participants believed their pharmacy services could better appreciate a person’s culture when providing care. One pharmacy technician stated *“I think people are polite (pause), generally. But I don’t think there’s anything in place to first recognise a patient’s cultural needs and then go from there”* (Participant 8).

Approaches that could improve the inclusivity of pharmacy services were discussed, with many relating to language and the need for translators or interpreters to support conversations about medicines. Translation and interpreter services were recognised as a potentially beneficial pharmacy service for patients who do not speak English as *“a lot of information that we give across to patients i*.*e*., *patient information leaflets, which are inside and the boxes and everything, well the majority are just in English”* (Participant 7). Another participant, who worked in an area with a higher level of deprivation, discussed strategies employed in their pharmacy which was managed by a pharmacist from an ethnic minority background. They described how steps towards being more culturally inclusive included *“translated medicine instructions on the wall sometimes posters are in different languages… I don’t know it just feels different working in that pharmacy compared to your average pharmacy, in that they’re more inclusive”* (Participant 9). It was also acknowledged by several participants that community pharmacy records *“may not have someone’s ethnicity in the notes that you can see”* (Participant 8) and the benefit of making this information readily accessible was suggested, to inform decisions made by pharmacy professionals about the best course of treatment for a patient.

> *“I’d say we could probably do more, and that’s probably all community pharmacies in general. And it’s even things like having leaflets in a different language available – I mean we can print them off, but do we? I’m aware of a company that’s er that does prescription transcribing in hundreds of different languages and it’s more accurate that Google Translate. But it’s only a very small number of pharmacies that are using it. But I think the ones that are, it is just so helpful to the patients” (Participant 10)*

#### Gaining trust and forming culturally-respectful relationships

Participants reflected on factors that might negatively impact on building culturally-respectful relationships; for example, the effect of language barriers if a person has limited or low English language proficiency, or use of colloquialisms when conversing with someone that does not speak English as a first language. A counter assistant reflected *“where I work you sort of know a lot of the people that come in – they’re like locals so you can have that like joke or bit of a chat with them. And with someone of a different background, you don’t always have that. And er, it can be a bit tricky”* (Participant 3). In fact, using cultural colloquialisms to build a better rapport with patients was highlighted by a pharmacist. They stated that within Asian cultures *“they call like ‘Auntie’ and ‘Uncle’, they use that language and it’s friendly language instead of ‘Mr or Mrs’. Er, yeah, very friendly terms used to create more of that rapport within the community”* (Participant 9).

Building trust and establishing culturally-respectful relationships was deemed by one participant to be potentially more challenging to achieve between a pharmacist and a patient from a different ethnicity or different culture. The employment of staff members from ethnic minority backgrounds was recognised as a strategy to overcome this. Many participants reflected on examples they had witnessed where staff members who spoke languages other than English were asked to speak to certain patients. One pharmacist stated that their pharmacy had *“quite a few Romanian customers who come in because they know that they can speak to someone in their language. Er, and I think they tell their friends, ‘If you go to this pharmacy, they’ll speak Romanian’”* (Participant 1). It was speculated that this approach could help overcome communication barriers between pharmacy staff and patients, by enabling inclusive conversations about medicines in a person’s native language. Further, it was considered that diversity within pharmacy staff may lead to greater patient satisfaction rates specifically focused on medicines; one participant stated *“so, I know it’s important to build a rapport up with someone, but then to be able to have a proper focused conversation understand it from a certain cultural perspective*… *that’s like, that’s achieving a proper goal for us”* (Participant 9). A dispensing assistant echoed this thinking, describing *“when a patient who comes in who’s a Sikh, for example, our pharmacist is a Sikh so he can focus on their background and kind of knows where they’re coming from when they’re asking about if it (their medication) is suitable”* (Participant 2).

### Theme 3: Educating and training cultural competence within the pharmacy profession

#### Perspectives on current cultural competence training

There was a consensus amongst participants involved in this study that their education and training was lacking in topics encompassing cultural competence and its place within the pharmacy profession. Some pharmacist participants stated that they remembered receiving some cultural competence training once they had qualified, but this was included as part of a *“bigger topic such as Equality & Diversity or Protected Characteristics”* within the workplace environment (Participant 7). These perspectives around the paucity of cultural competence training were shared with other community pharmacy staff, where one counter assistant stated that *“you just sort of have to deal with it yourself, you don’t really get any training on that”* (Participant 3). In fact, there was a common opinion that due to the lack of cultural competence training within professional teaching, it was necessary for individuals to educate themselves on cultural competence. One pharmacist reported *“this thinking wasn’t built into our teaching really. It wasn’t er something we were taught; it was just a good thing for you to look up in your spare time”* (Participant 1).

All interviewees regarded cultural competence as a necessary and important skill and all were in agreement that some form of education should be built into pharmacy staff training. One pharmacy technician stated *“I think whoever you are, if you are talking to members of the public who come into the pharmacy or are someone who speaks to patients, you should have gone through a minimum level of training on cultural competence”* (Participant 8). However, across the different job roles within community pharmacy, there was a mixed consensus on who should receive cultural competence training and to what level/depth. Almost all participants felt that cultural competence training should be provided for counter assistants, with one participant stating *“a person walks in and the first people they’ll speak to is the counter staff”* (Participant 5). It was acknowledged that if counter assistants were not trained on cultural competence, *“it may affect the treatment of a patient and could potentially impact how much a patient will use the pharmacy or whether they come back or feel comfortable even coming in(to the pharmacy)”* (Participant 1). When discussing their perspectives, a pharmacy dispenser expressed that pharmacists *“are the main people who are giving advice out so if they’re not competent in what they’re saying (pause), then it’s not going to help the situation”* (Participant 2).

Despite acknowledging the importance and place of cultural competence training, a number of participants suggested that the profession of pharmacy, as a whole, has unmet needs around the training and delivery of culturally competent patient care. One participant mentioned that, following the Black Lives Matter movement, they have requested additional training on cultural competence from their employer to improve their knowledge. Another reflected on the growing discussion of equality and inclusivity in wider society, stating that *“the (COVID-19) pandemic was what brought the whole issue about inequalities to light in my mind, and since then I’ve been thinking about what it means for these patients from other communities”* (Participant 13).

> *“… I don’t think, personally, as a profession, we’re prepared really. And we don’t have that awareness – I certainly don’t feel like I’m optimally prepared for dealing with situations where I have a lack of knowledge about someone’s ethnic background. It’s actually something that I’ve requested at work is some training around it” (Participant 5)*

#### Suggestions for the delivery of cultural competence training

Participants gave suggestions on how cultural competence could be incorporated into formalised training for each professional group. The training for counter assistants was described involving *“booklets at the start… which you had to complete, and you had so long to complete them. And if you pass them, you’re classed as a counter assistant”* (Participant 3). The same participant went on to mention that *“it [cultural competence training] should be in the booklet when you initially start, and that should definitely be covered in the booklet”* (Participant 3). Another counter assistant held a different opinion and suggested that *“if it was in a training handbook like where you watch videos and answer quizzes, then people maybe wouldn’t take it seriously. Er, like people might do the quiz and not actually care or take it in as actually being important”* (Participant 12). They reflected on the potential for variation in the population of each pharmacy, dependent on geographic location, as well as postulating a constant, dynamic change in *“people living nearby, ‘cos those trends of communities choosing to live in a certain area might change”* (Participant 12). When asked about what should be taught within this training, it was proposed that a basic understanding and awareness of different cultures should be taught *“I think just explaining what it (cultural competence) is and maybe an idea on how different backgrounds, you know, different people – their views on sort of… pharmacy and medicines and things like that”* (Participant 3).

There was a suggestion made that it may be beneficial to teach cultural competence training separately within the different professions and tailoring the training towards each professions’ specific roles within the pharmacy. A dispenser proposed that *“if everyone’s taught separately but on the same kind of guidelines, so it (cultural competence training) kind of factors in their different job roles… then it’s helpful if it’s got some examples of what we can do, personally, relevant to us”* (Participant 2).

Every pharmacist interviewed agreed that cultural competence training should be taught within the undergraduate Masters of Pharmacy (MPharm) degree; however, there were differing perspectives on the value it holds within the degree’s curriculum. One pharmacist remarked *“I think it should be taught, I think it’s core, it’s just as important as learning about pregnancy and drugs in pregnancy… it needs to be included in the curriculum”* (Participant 5). As well, participants discussed the need for cultural competence to underpin the entirety of their learning, rather than only featuring in one part of the degree. There was a particular emphasis on introducing *“this way of thinking”* from the first year of the degree, with one foundation trainee pharmacist discussing the value in having *“these types of discussions or training from like an earlier stage, and from a younger age at university*… *that would be good as opposed to leaving it until you’re in the workplace like I am now”* (Participant 14). Whilst other pharmacists recognised its importance, they considered the logistics of integrating cultural competence training within the greater context of the MPharm degree curriculum and acknowledged that there may limited room to include this training. One pharmacist stated that cultural competence training should be *“definitely part of it [the MPharm]. I don’t know if I’d say a big part. Er (pause) not because it’s not important – just thinking more in terms of how you’d fit it all in”* (Participant 6).

Pharmacists offered suggestions on how cultural competence training could be delivered within the MPharm degree. The most frequently mentioned methods considered adopting an integrated approach to teaching, for example through the inclusion of ethnic minority patients within workshops, patient-case examples and observed structured clinical examination (OSCE) stations, as well as exposing students to working with people from ethnic minority communities on placements.

> *“In case studies when you’re doing seminars, you should ensure there’s varied names and ethnicities used in patient examples. Perhaps trying to encompass it more into placements as well, where you are actually in real-life examples, so you’ve got real-life patients and are seeing varied cross-sections of the population in placements. You could potentially look at patient sessions where you bring patients in and getting different backgrounds to talk about their experiences” (Participant 7)*.

Two participants contemplated that, rather than training students on cultural competence, the aim of the MPharm teaching should be to instil an open-minded and inquisitive attitude in students for whenever they engage with people from ethnic minority groups. These participants argued that giving cultural training would not be appropriate, with one stating *“to sit down and have a lesson and say, ‘This is how you speak to someone who’s Muslim’ or ‘This is how you speak to someone who is from this ethnicity’ – I think that’s patronising”* (Participant 1).

> *“I think it’s really important that students don’t become stereotypical and judge people and assume based on the way that someone looks, that they have a certain belief. I think it’s more about students being open-minded (pause) and having the ability to ask these questions*… *I just feel that’s a skill we need to get rather than understanding all the nuances of different cultures cos you can’t teach that – it’s not possible. It’s more about giving them the skills and then once they work in practice, they deal with the communities that they encounter” (Participant 4)*

## Discussion

This study builds on the limited evidence that focuses on the perspectives of community pharmacy staff surrounding cultural competence training. By exploring the perspectives of members of staff across the entire skill mix of community pharmacy, this study (i) sheds new light on staff perspectives on cultural competence training and (ii) offers unique suggestions on how cultural competence training should be taught and delivered to all members of the community pharmacy team. This study collected the perspectives from representatives of all community pharmacy staff members: pharmacists, technicians, dispensers and counter assistant staff; an approach that is unique to this study with these views previously being under-reported within healthcare research.

A consistent finding across all interviews reported an increase in the ethnic diversity of people accessing community pharmacies, hence recognising the greater need for community pharmacy staff to adopt an inclusive approach towards meeting the needs of their ethnic minority patient groups. This echoes the findings from previous work^11,23^ and has been highlighted even more following the COVID-19 pandemic.^33-35^ Ethnicity-related health inequalities is a well-researched area, with numerous studies reporting a lack of patient engagement with services and variable treatment outcomes in patients from ethnic minority groups, compared to their White counterparts. Multifactorial reasons for this have been recognised, including perceived distrust in the healthcare system, lower health literacy, limited access to timely care and language barriers;^12,36-38^ initial strategies to overcome such challenges, specifically within community pharmacy, have been identified in this study.

Echoing previous studies,^39,40^ communication difficulties and language barriers between patients and healthcare professionals were perceived as a key challenge in achieving inclusive patient care within community pharmacy settings. Interestingly, encounters with non-English speaking patients were also a major concern amongst other healthcare professional groups.^41^ Previous work revealed that computer-based translational resources were perceived as easily accessible and helpful within American community pharmacy, but were infrequently used.^42-44^ “The Written Medicine,” is a new UK web-based software which has been developed to tackle this issue through bilingual prescription labels, including English alongside the person’s native language.^45^ Future research should explore the uptake and utilisation of such software within community pharmacies.

The use of interpreters was also a recognised approach to overcoming communication difficulties between patients with Low-English Proficiency and members of the pharmacy team. Interpreters were also important for providing high quality care to these patients. Although the use of professional interpreters has been acknowledged as beneficial to facilitate healthcare consultations within outpatient clinical settings,^46-49^ there has been a longstanding historical challenge to accessing interpreters within community pharmacy.^50-52^ Research on facilitating access to interpreters and exploring pharmacy team experiences with interpreters could result in the sharing of best practice, to improve culturally competent and medicines-focused communication.

Whilst participants recognised the significance of cultural competence training, they reported very little to no experience of having cultural competence training within their professional education. Results from this study echo the wider literature,^53,54^ which recognises the importance of integrating cultural competency into the training of pharmacy staff. In 2021, the Centre for Pharmacy Postgraduate Education launched a learning campaign on ‘Culturally competent person-centred care’, designed for pharmacy professionals.^55^ At present, research has yet to be done to evaluate the impact or uptake of this amongst postgraduate, qualified pharmacists. Further, limited work exists to explore the perspectives of student and trainee pharmacists on cultural competence within their initial education and training; future studies should seek to address this to close the gap between training at undergraduate and postgraduate level.

A key relationship was identified between participant ethnicity and their views on the importance of cultural competency, with its training being deemed important by all White participants who were interviewed. However, two participants who self-reported as being from an ethnic minority group themselves (from Chinese and Pakistani backgrounds), hinted that cultural competence training does not necessarily need to be a key part of professional training. Several White participants stressed the importance of this training as, within their practice, they often worry about unintentionally using the incorrect language/terminology.^56,57^ This study offers a new finding that White healthcare professionals may regard cultural competence training as more important than those from ethnic minority backgrounds; further research is needed to explore whether this is the case and identify reasons why this may exist.

The researcher acknowledges that there were some limitations in this study. Whilst efforts were taken to ensure an equal split in job roles amongst the participants, the majority were pharmacists (n=8). Future studies should seek to further explore the perspectives of the wider roles within community pharmacy teams. The most common ethnicity among the participants was White, British; in order to make this study more representative, the perspectives of staff members from underrepresented ethnic communities need further investigation. In doing so, this may help to determine if culture plays a part in how cultural competence, and training, is perceived.

Perspectives of community pharmacy staff have been reported within this study. Future studies could investigate the perspectives of underrepresented ethnic patient groups on how they believe cultural competence training should be delivered to community pharmacy staff, to allow a cooperative approach to re-evaluating the training strategies.^58^ Previous studies have reviewed co-production and co-design approaches to tailor health services to the needs of its users.^59,60^ Therefore, including the perspectives of people from underrepresented ethnic groups would allow their lived experiences and suggestions to be considered when formulating and reviewing cultural competence training to community pharmacy staff. Furthermore, research could be widened even more, by exploring perspectives on how cultural competence teaching can be promoted to address the needs of other minoritised patient groups.

## Researcher positionality and reflexivity statement

When conducting research on cultural competence and its wider connection to ethnicity, it is important to acknowledge the positionality and reflexivity of the research team. Authors JMcC, AH, AT and AR-B recognized their privilege as nonethnic minority UK citizens. Author WML, and patient champions LS and AKD self-report as being from an ethnic minority community; they ensured cultural appropriateness and sensitivity throughout the entire research process.

## Conclusion

Cultural competence is important for community pharmacy staff to develop, due to the increasing patient diversity they encounter. As pharmacists and counter assistants are primarily patient-facing roles, cultural competence training should be an essential part of their professional training. Interactive, informal teaching to encourage an open-minded attitude towards cultural differences, was preferred over structured, information-based sessions. Future research may seek to further explore the integration of cultural competence training with undergraduate pharmacist initial education and training, as well as postgraduate learning programmes. Research opportunities into co-production and co-design strategies between trainees and training organisations could improve the design of cultural competence training. Additionally, the perspectives of patients from ethnic minority communities could provide valuable insight and offer recommendations on the cultural competence training delivered to pharmacy professionals, and wider.

## Data Availability

All data produced in the present work are contained in the manuscript.
Further data that support the findings of this study are available from the corresponding author upon reasonable request.

## Acknowledgements

The authors would like to thank all of the participants involved in this research; without whom, this work would not be possible. The authors would also like to thank the wider steering group patient champions who made this work possible: Manpreet Olk, Evneesh Dhinsa, Samia Fiazei and Jel Nagra.

The authors wish to acknowledge and thank the participants that were interviewed as part of this research; without whom, this work would not have been possible.

## Data availability statement

The data that support the findings of this study are available from the corresponding author upon reasonable request.

## Author contributions

JMcC led on the day-to-day running of the project, data collection and writing of this manuscript. AR-B and WML oversaw the running of this project as Principal Investigators and provided project management expertise. AR-B, AH and AT provided qualitative methodological input and expertise. AR-B and AH supported the recruitment of participants. LS, SV and AKD contributed in their appointment as ethnicity champions and ensured cultural appropriateness and sensitivity throughout the entire research process. All authors read, provided comments on, and approved the final manuscript.

## Ethics approval statement

Ethical approval was obtained from the [anonymised] University Research and Ethics Committee (reference: 25680/2022).

## Patient consent statement

There were no patients involved in this study; it was conducted to gather the perspectives of healthcare professionals (and signed consent was gained at study enrolment). Permission to reproduce material from other sources: Not applicable; this work is original.

## Clinical trial registration

Not applicable.

