## Supplementary File for "“*Creating a culturally competent pharmacy profession*”: a qualitative exploration of pharmacy staff perspectives of cultural competence and its training in community pharmacy settings"

**Item 1:**

**COnsolidated criteria for REporting Qualitative studies (COREQ): 32-item checklist.**

| **Number** | **Item** | **Guide questions / description** | **Reported on manuscript page** |
| --- | --- | --- | --- |
| **Domain 1: research team and reflexivity** | | | |
| **Personal characteristics** | | | |
| 1 | Interviewer | Which author(s) conducted the interviews? | 6 |
| 2 | Credentials | What were the researcher’s credentials? *E.g., PhD, MD* | 6 |
| 3 | Occupation | What was their occupation at the time of the study? | 6 |
| 4 | Gender | Was the researcher male or female? | 6 |
| 5 | Experience and training | What experience or training did the researcher have? | 6 |
| **Relationship with participants** | | | |
| 6 | Relationship established | Was a relationship established prior to study commencement? | 5 |
| 7 | Participant knowledge of interviewer | What did the participants know about the researcher?  *E.g., reason for doing the research* | 5 |
| 8 | Interviewer characteristics | What characteristics were reported about the interviewer?  *E.g., bias, assumptions, reasons and interests in the research topic* | 6 |
| **Domain 2: study design** | | | |
| **Theoretical framework** | | | |
| 9 | Methodological orientation and theory | What methodological orientation was stated to underpin the study?  *E.g., grounded theory, ethnography, discourse analysis* | N/A |
| **Participant selection** | | | |
| 10 | Sampling | How were participants selected? *E.g., purposive, convenience, consecutive* | 5 |
| 11 | Method of approach | How were participants approached? *E.g., face-to-face, telephone, email* | 5 |
| 12 | Sample size | How many participants were in the study? | 8, Table 1 |
| 13 | Non-participation | How many people refused to participate or dropped out (with reasons)? | 8 |
| **Setting** | | | |
| 14 | Setting of data collection | How was the data collected? *E.g., home, clinic, workplace* | 5, 8, Table 1 |
| 15 | Presence of non-participants | Was anyone else present besides the participant and researcher? | N/A |
| 16 | Description of sample | What are the important characteristics of the sample? *E.g., demographic data* | 8, Table 1 |
| **Data collection** | | | |
| 17 | Interview guide | Were questions and prompts provided by the authors? | 6, Supplementary file |
| 18 | Repeat interviews | Were repeat interviews carried out? If yes, how many? | N/A |
| 19 | Audio/visual recording | Did the researcher use audio or visual recording to collect the data? | 6 |
| 20 | Field notes | Were field notes made during/after the interview? | 6-7 |
| 21 | Duration | What was the duration of the interviews? | Table 1 |
| 22 | Data saturation | Was data saturation discussed? | 7 |
| 23 | Transcripts returned | Were transcripts returned to participants for comment/correction? | 6 |
| **Domain 3: analysis and findings** | | | |
| **Data analysis** | | | |
| 24 | Number of data coders | How many data coders coded the data? | 6 |
| 25 | Description of the coding tree | Did authors provide a description of the coding tree? | N/A |
| 26 | Derivation of themes | Were themes identified in advance or derived from the data? | 7 |
| 27 | Software | What software, if applicable, was used to manage the data? | 7 |
| 28 | Participant checking | Did participants provide feedback on the findings? | 7 |
| **Reporting** | | | |
| 29 | Quotations presented | Were participant quotations presented to illustrate the themes / findings? Was each quotation identified? E*.g., participant number* | 11-21 |
| 30 | Data and findings consistent | Was there consistency between the data presented and the findings? | 11-21 |
| 31 | Clarity of major themes | Were major themes clearly presented in the findings? | 11-21, Figure 1 |
| 32 | Clarity of minor themes | Is there a description of diverse cases or discussion of minor themes? | 11-21, Figure 1 |

**Item 2:**

**Semi-structured interview topic guide**

The semi-structured interview questions were based around the following topic areas:

1. (Broader) Experiences of cultural and ethnicity health in general
2. (Broad) Experiences of cultural and ethnicity within their personal life / place of work
3. (Narrowing) Experiences of engaging with cultural competence
4. (Narrow) Participant understanding of cultural competence
5. (Focused) Examples of inequalities within pharmacy profession / within place of work
6. (Focused) Interacting with patients from another ethnicity
7. (Focused) Training around cultural competence – current / previous
8. (Focused) Suggestions on supporting education on cultural competence
